# Temporary hold of mycophenolate boosts SARS-CoV-2 vaccination-specific humoral and cellular immunity in kidney transplant recipients

**DOI:** 10.1101/2022.01.05.21268478

**Authors:** Eva Schrezenmeier, Hector Rincon-Arevalo, Annika Jens, Ana-Luisa Stefanski, Charlotte Hammett, Bilgin Osmanodja, Nadine Koch, Bianca Zukunft, Julia Beck, Michael Oellerich, Vanessa Pross, Carolin Stahl, Mira Choi, Friederike Bachmann, Lutz Liefeldt, Petra Glander, Ekkehard Schütz, Kirsten Bornemann-Kolatzki, Covadonga López del Moral, Hubert Schrezenmeier, Carolin Ludwig, Bernd Jahrsdörfer, Kai-Uwe Eckardt, Nils Lachmann, Katja Kotsch, Thomas Dörner, Fabian Halleck, Arne Sattler, Klemens Budde

**Affiliations:** Department of Nephrology and Medical Intensive Care, Charité-Universitätsmedizin Berlin, corporate member of Freie Universität Berlin and Humboldt-Universität zu Berlin, Berlin, Germany; Department of Rheumatology and Clinical Immunology, Charité-Universitätsmedizin Berlin, corporate member of Freie Universität Berlin and Humboldt-Universität zu Berlin, Berlin, Germany; Deutsches Rheumaforschungszentrum (DRFZ), Berlin, Germany; Grupo de Inmunología Celular e Inmunogenética, Facultad de Medicina, Instituto de Investigaciones Médicas, Universidad de Antioquia UdeA, Medellín, Colombia; Department of Clinical Pharmacology, University Medical Center Göttingen, Göttingen, Germany; Department for General and Visceral Surgery, Charité-Universitätsmedizin Berlin, corporate member of Freie Universität Berlin and Humboldt-Universität zu Berlin, Berlin, Germany; Chronix Biomedical GmbH, Göttingen, Germany; Institute of Transfusion Medicine, Ulm University, Ulm, Germany and Institute for Clinical Transfusion Medicine and Immunogenetics. German Red Cross Blood Transfusion Service Baden-Württemberg – Hessen and University Hospital Ulm, Ulm, Germany; Center for Tumor Medicine, H&I Laboratory, Charité University Medicine Berlin, Berlin, Germany; Berlin Institute of Health at Charité–Universitätsmedizin Berlin, BIH Academy, Clinician Scientist Program Universitätsmedizin Berlin, Berlin, Germany

## Abstract

Transplant recipients exhibit an impaired protective immunity after SARS-CoV-2 vaccination, potentially caused by mycophenolate (MPA) immunosuppression. Recent data from autoimmune patients suggest that temporary MPA hold might significantly improve booster vaccination outcomes. We applied a fourth dose of SARS-CoV-2 vaccine during temporary (5 weeks) MPA hold to 29 kidney transplant recipients, who had not mounted a humoral immune-response to previous vaccinations. Seroconversion until day 32 after vaccination was observed in 76% of patients, associated with acquisition of virus neutralizing capacity. Interestingly, 21/25 (84%) CNI-treated patients responded, but only 1/4 Belatacept-treated patients. In line with humoral responses, counts and relative frequencies of spike receptor binding domain (RBD) specific B cells were significantly increased on day 7 after vaccination, with an increase in RBD specific CD27^++^CD38^+^ plasmablasts. Whereas overall proportions of spike-reactive CD4^+^ T cells remained unaltered after the fourth dose, frequencies were positively correlated with specific IgG levels. Importantly, antigen-specific proliferating Ki67^+^ and in vivo activated PD1^+^ T cells significantly increased after re-vaccination during MPA hold, whereas cytokine production and memory differentiation remained unaffected. In summary, MPA hold was safe and augmented all arms of immunity during booster vaccination, suggesting its implementation in vaccination protocols for clinically stable transplant recipients.

## Introduction

Protection of kidney transplant recipients (KTR) from coronavirus disease 2019 (COVID-19), caused by severe acute respiratory syndrome coronavirus 2 (SARS-CoV-2), has not been sufficiently achieved by conventional vaccination protocols. Mortality of fully vaccinated KTR after infection remains unacceptably high with almost 8% in a registry analysis from the United Kingdom (1) and up to 20% in other cohorts (2, 3), despite the presence of vaccine-specific T cells. Our previous studies revealed a strong impairment of both humoral and cellular immunity in transplant recipients after two doses of BNT162b2, with antigen-specific B and T cell responses being both quantitatively and functionally affected (4, 5). Since the majority of KTR does not benefit from a third dose (6), modified vaccination protocols are required to achieve protection of this at-risk population.

Analysis of larger transplant patient cohorts indicated that mycophenolate (MPA) based treatment constitutes a major risk factor for impairment of vaccine-induced humoral immunity (7, 8). In line with the aforementioned, a case series with patients suffering from rheumatic and musculoskeletal diseases demonstrated that temporary hold of MPA leads to augmented humoral responses to SARS-CoV-2 vaccination (9). According to European guidelines, the majority of kidney transplanted individuals receive triple immunosuppressive medication including calcineurin inhibitors (CNI), corticosteroids (CS) and MPA. Withdrawal of steroids or MPA in a tacrolimus-based treatment protocol for up to three years has been shown to be safe in a large multicenter study, with no increase in acute rejections or impaired kidney function (10). Similar data were obtained from other trials (11-13). Furthermore, hold of MPA is routinely recommended during pregnancy (14), underlining the feasibility of this approach. To examine the impact of short-term MPA withdrawal on vaccination outcome, 29 KTR, being seronegative after triple SARS-CoV-2 vaccination, were converted to an MPA-free immunosuppressive regimen. Patients were closely monitored for clinical parameters, including kidney function, anti-human leucocyte antigen (HLA) antibodies and donor-derived cell-free DNA (dd-cfDNA) (15, 16); assessment of vaccine-specific immunity encompassed in-depth analysis of specific B and T cell analyses, IgG and IgA levels and neutralization capacity.

## Results

### Vaccination-induced humoral and B-cell immunity

The study cohort included 29 KTR with a lack of serological response after a three-dose vaccine protocol. Fourteen patients were homogeneously vaccinated (3x mRNA vaccine), 15 patients were vaccinated heterologously (mixed mRNA- and vector-based). All patients received BNT162b2 (BioNTech/Pfizer) as a fourth vaccine. Mean time interval between the third and fourth vaccination was 59.1 (±12.6) days. All patients were initially on antimetabolite treatment, 28/29 on MPA and 1/29 on azathioprine (Aza). 25/29 received CNI based medication while 4 patients received Belatacept. All patients stopped MPA or Aza 4-7 days before the fourth vaccination, based on the assumption that pharmacodynamic drug effects wean after 3-4 days (17). Treatment was paused until day 28-35 (mean depicted as “day 32” in all figures; second time point for serological response analysis). In patients with no or low corticosteroid (CS) medication, CS was restarted or increased to 5 mg prednisone equivalent together with MPA hold. In the four patients on Belatacept, two stopped MPA and one Aza, while it was replaced by CS in only one patient. One Belatacept treated patient was switched to tacrolimus and CS. Demographics are summarized in Table 1.

**Table 1:** Patient demographics.

| **Table I** |  |
| --- | --- |
| Characteristics of kidney transplant (KTX) patients enrolled (n=29) |  |
| **Variable** |  |
| Age (mean yrs ± SD) | 59.8 (14.8) |
| Females (%) | 12 (41.4) |
| Males (%) | 17 (58.6) |
| Caucasians (%) | 29 (100) |
| Homologous vaccine protocol (%) | 14 (48.3) |
| Heterologous vaccine protocol (%) | 15 (51.7) |
| Time (days) between the third and fourth vaccination (SD) | 59.1 (±12.6) |
| **Clinical Parameters** |  |
| Time since Tx (mean yrs ± SD) | 9.9 (5.9) |
| Retransplantation (%) | 3 (10.3) |
| Living donor transplantation (%) / ABOi (%) | 15 (51.7) / 4 (26.7) |
| Acute graft rejection after 4<sup>th</sup> vaccine (%)<sup>A</sup> | 0 (0) |
| **Immunosuppressive medication before change** |  |
| Tac+MPA (%) | 4 (13.8) |
| CS+Tac+MPA (%) | 14 (51.7) |
| CS+CyA+MPA (%) | 6 (20.7) |
| CyA+MPA (%) | 1 (3.4) |
| Belatacept+Aza ±CS (%) | 1 (3.4) |
| Belatacept+MPA ±CS (%) | 2 (6.8) |
| Belatacept+MPA (SD) | 1 (3.4) |
| Mean MPA equivalent dose | 1.2 (0.7) |
| **Laboratory parameters before 4<sup>th</sup> vaccination** |  |
| Mean eGFR ml/min (SD) | 45.4 (14.8) |
| Mean IMPDH activity (pmol XMP/h/mg Hb) (SD) | 1192.7 (474.2) |
| **Laboratory parameters after 4<sup>th</sup> vaccination 1-3 months** |  |
| Mean eGFR ml/min (SD) | 48.6 (19.7) |
| **Comorbidities** |  |
| Hypertension (%) | 24 (82.3) |
| Coronary heart disease (%) | 6 (20.6) |
| Diabetes (%) | 2 (6.9) |
| History of malignancy (%) | 1 (3.4) |
| CS – Corticosteroids, Tac – Tacrolimus, CyA – Cyclosporin A, MPA – Mycophenolate, ABOi- ABO incompatible living kidney donation. SD – standard deviation. <sup>A</sup>between MPA hold and day 32 |  |

Seroconversion (OD ratio>1.1) for anti-S1 domain IgG occurred in 10/29 (34.5%) individuals until day 7 after the fourth vaccination, while anti-S1 domain IgA was positive in 7/29 KTR (24.1%). Neutralization capacity above 30% was achieved in 11/29 (37.9%) patients (Figure 1A-C). On day 32 after vaccination, 22/29 (76%) patients showed anti-S1 domain IgG levels above the threshold for positivity. Anti-S1 domain IgA and neutralization capacity levels were unavailable for 8 individuals. For the remaining individuals, IgA was positive in 11/21 patients (52.2%) and neutralization capacity was above threshold in 15/21 (71.4%) patients (Figure 1A-C). In patients with CNI treatment before vaccination, anti-S1 domain IgG seroconversion occurred in 21/25 (84%) patients, while 3/4 patients on Belatacept remained negative; only one became weakly positive just above the threshold on day 32 (Supplementary Figure 1B). Anti-S1 domain specific IgG on day 32 did not differ between patients upon heterologous or homologous vaccination (Supplementary Figure 1A).

**Figure 1:**
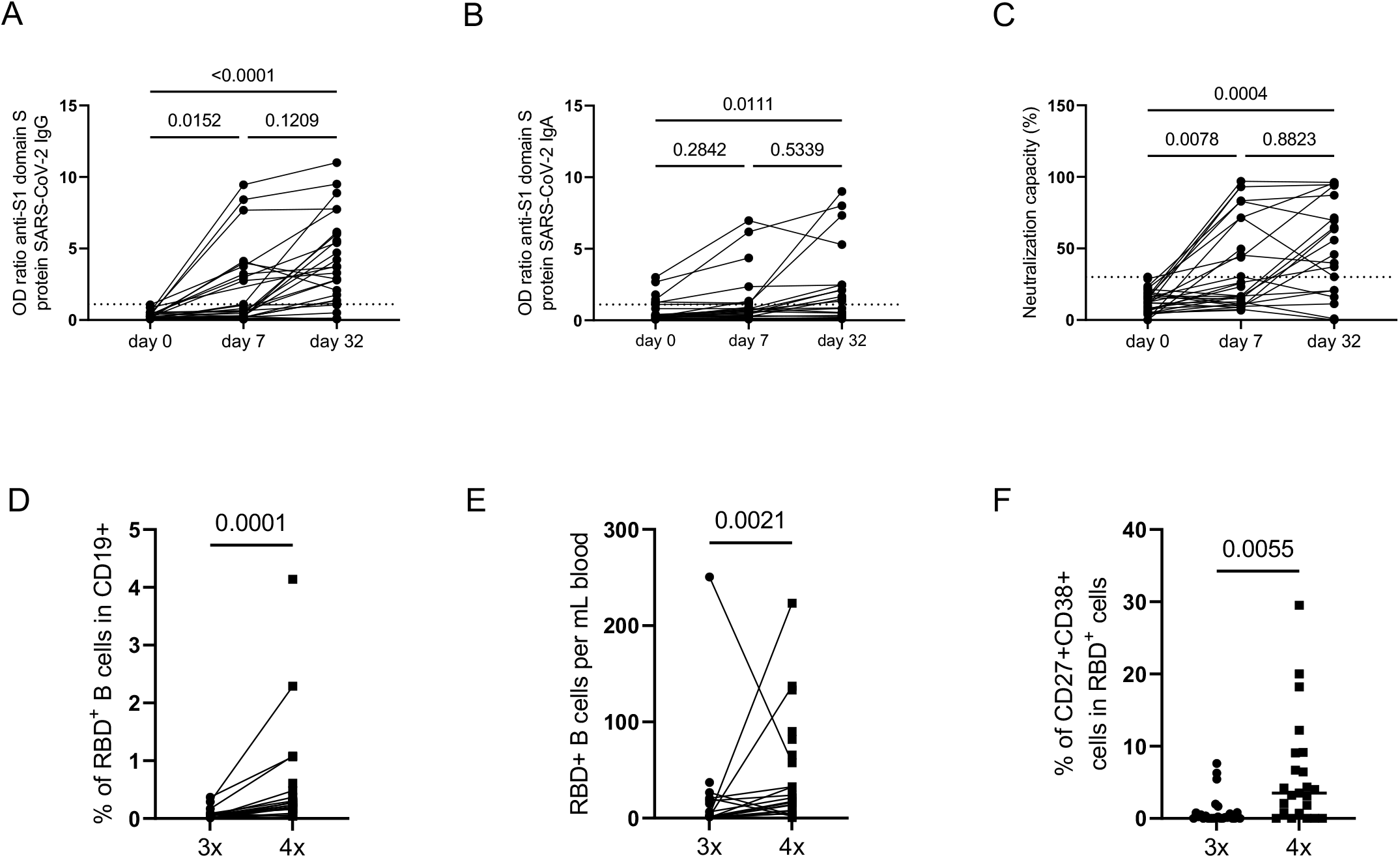
Humoral immune responses and specific B cell immunity after fourth vaccination in KTR. Humoral vaccine-specific immune responses were assessed by ELISA for anti-spike protein S1 IgG (n=29/29) (A), spike protein S1 IgA (29/29 day 7, 21/29 day 32) (B) and virus neutralization by a blocking ELISA (n=29/29 day 7, n=21/29 day 32) (C) at the indicated timepoints in KTR after administration of a fourth dose BNT162b2. Thresholds defining a positive response are indicated by dotted lines. (A) Relative frequencies (3x – n=25, 4x – n=23) and (B) absolute counts (3x – n=23, 4x – n= 23) of RBD-specific CD19^+^ B cells 7±2 days after fourth vaccination with BNT162b2. (C) Frequency of RBD-specific CD27^++^CD38^+^ plasmablasts. (A-C) Kruskal-Wallis with Dunn’s post-test. (D, E, F) Mann-Whitney test. Where applicable, graphs show means ± SD.

Antigen-specific B cells were identified by fluorescent double-labelling of reactive cells with recombinant receptor binding domain (RBD) (5) (Supplementary Figure 1D). Frequencies and absolute counts of RBD^+^ B cells increased 7 days after vaccination compared to baseline (Figure 1D). Interestingly, especially the frequency of RBD^+^ plasmablasts, which have been shown to be an early sign of vaccine response (5), increased after four vaccinations (Figure 1E).

The inosine monophosphate dehydrogenase (IMPDH) activity in erythrocytes has recently been described as a useful pharmacodynamic marker for MPA exposure that reflects the MPA exposure after 8 weeks of constant dosing (17). High IMPDH levels were found in patients with MPA toxicity and low levels in patients with biopsy-proven acute rejections (17). In the current cohort, the mean IMPDH activity before MPA hold at a time of steady state was 1192.73 pmol XMP/h/mg Hb (±474.24), and IMPDH activity did not negatively correlate with anti-S1 domain IgG on day 32 (Supplementary Figure 1C).

### Vaccination-specific CD4^+^ T cell responses

SARS-CoV-2 spike protein reactive CD4^+^ T helper cells were detected within PBMC based on activation-induced co-expression of CD154 and CD137 after stimulation with 15-mer peptides (overlapping by 11 amino acids, respectively) covering the complete spike glycoprotein sequence, as previously reported (4, 18). The gating strategy, including subset identification, is depicted in Supplementary Figure 2. A positive T cell response was defined when stimulated PBMC contained more than threefold higher frequencies of CD154^+^CD137^+^ CD4^+^ T cells as compared to the unstimulated control (stimulation index of 3) with at least twenty events, being in accordance with comparable studies (19). The prevalence of cellular responders was similar (>85%) after the third and fourth vaccination with no significant differences in relative and absolute frequencies of antigen-reactive T cells. Of note, levels of anti-SARS-CoV-2 spike S1 domain specific IgG were positively correlated with frequencies of spike-specific T cells (Figure 2A).

**Figure 2:**
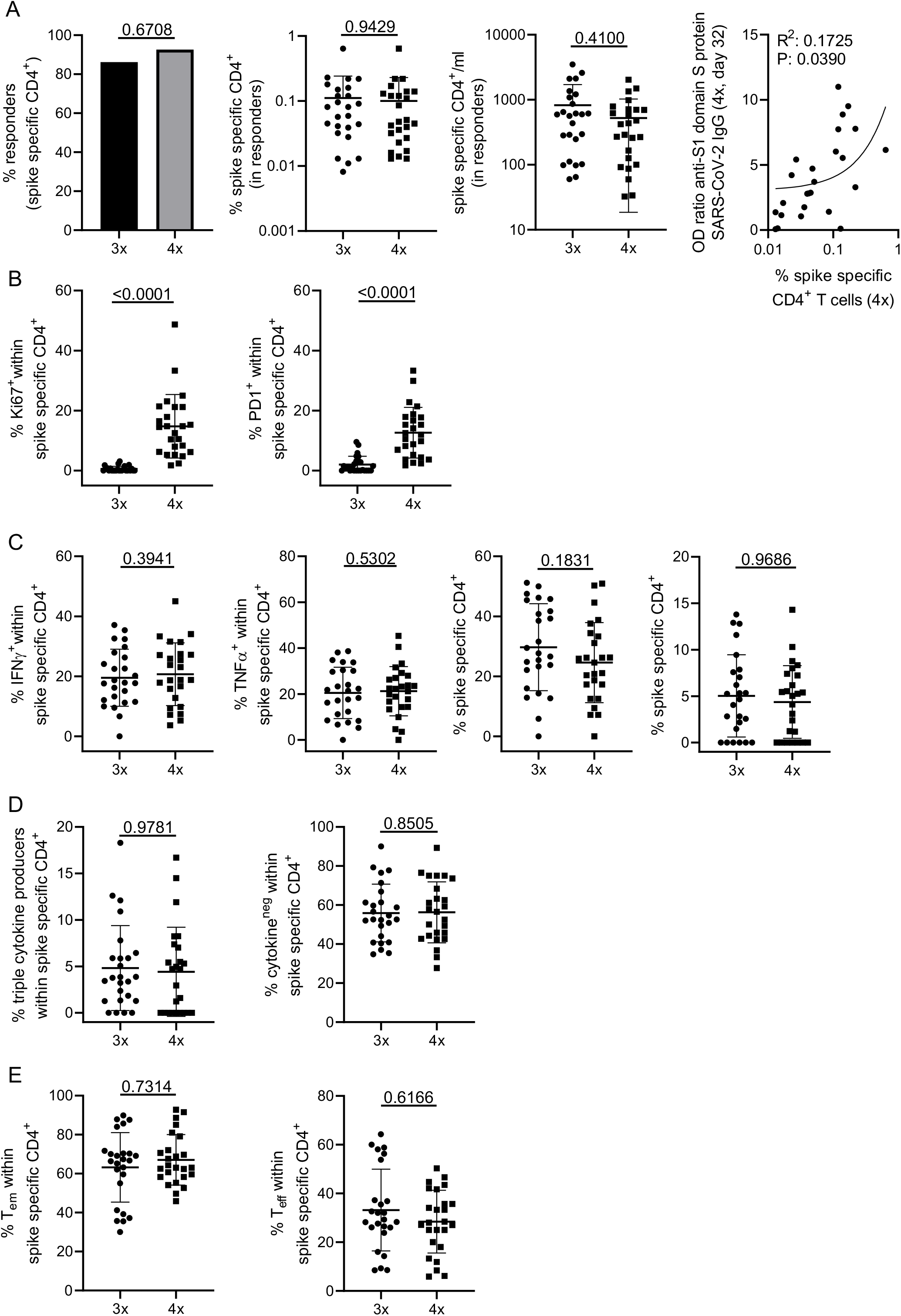
Assessment of T cell reactivity. PBMC of KTR were stimulated with spike peptide mix or left unstimulated. Specific CD4^+^ T cells were detected immediately before the fourth dose and 7 days thereafter by flow cytometry according to co-expression of CD154 and CD137. (A) depicts the portion of individuals with a cellular response (left, Fisher’s exact test, 3x-n=29, 4x-n=27), relative (middle/left, paired Wilcoxon test) and absolute (middle/right, paired Wilcoxon test) frequencies of specific CD4^+^ T cells and the correlation between relative frequencies and levels of anti-spike S1-domain IgG (right, simple linear regression). (B) Frequencies of antigen-reactive CD4^+^ T cells expressing Ki67 (left, paired Wilcoxon test) or PD1 (right, paired Wilcoxon test). (C) Expression of IFNγ (paired t test), TNFα (paired t test), IL-2 (paired t test) and IL-4 (paired t test) in antigen-specific T cells. (D) Analysis of IFNγ^+^TNFα^+^IL-2^+^ “triple^+^” polyfunctional (paired Wilcoxon test, left) and cytokine-non-producing cells (paired Wilcoxon test, right). (E) Memory/effector subset differentiation of antigen-specific CD4^+^ T cells (T_EM_: effector memory (left, paired Wilcoxon test), T_eff_: effector (right, paired Wilcoxon test). In all analyses except in responder rate calculation (A), n=25 individuals were included per group. Where applicable, graphs show means ± SD.

Spike-specific T cells of individuals after the fourth vaccination contained significantly higher portions of cells expressing the proliferation marker Ki67; the same applied to expression of PD-1, indicating recent in vivo activation (Figure 2B). Interestingly, we did not detect significantly elevated frequencies of antigen-specific T cells expressing IFNγ, TNFα, IL-2 or IL-4 after the fourth dose (Figure 2C); this also applied to proportions of specific polyfunctional IFNγ^+^TNFα^+^IL-2^+^ T cells or cells secreting none of the three cytokines (Figure 2D). IL-4 was excluded from polyfunctionality analyses due to low frequencies of positive cells. Antigen-reactive T cells from individuals after the third and fourth dose showed similar frequencies of CD45RO^+^CD62L^-^ effector/memory and CD45RO^-^CD62L^-^ effector-type T cells, respectively (Figure 2E).

### HLA antibody testing and dd-cfDNA

Since conversion of an established immunosuppressive regimen bears the risk of adverse events such as rejection or the generation of de-novo HLA antibodies, we performed anti-HLA antibody testing before and after vaccination in 27/29 study participants. No patient developed de-novo HLA antibodies, and antibody pattern and strength remained unchanged in 3 patients with preexisting donor-specific HLA antibodies before vaccination. Kidney function remained stable after day 32. Dd-cfDNA, a novel marker for subclinical allograft injury and rejection (16, 20) was available for 16/29 KTR and did not increase.

## Discussion

The current study investigates the immunological impact and clinical safety of a fourth dose of a SARS-CoV-2 vaccine during short-term MPA hold in KTR who did not seroconvert after three vaccine doses. We found a significant increase in humoral responders at day 32 (76% irrespective of previous treatment, 84% in individuals on standard CNI regimen), together with an increase in neutralizing antibodies and occurrences of vaccine-specific B cells and plasmablasts. We further observed higher ex vivo activation of spike-specific T cells that quantitatively correlate with spike S1 specific IgG at day 32.

So far, the response to SARS-CoV-2 mRNA- and vector-based vaccines in KTR has been disappointing (4, 21), resulting in high infection and hospitalization rates in fully vaccinated individuals (3). The early recommendation of a third vaccination for solid organ recipients, that has meanwhile been extended to the general population (22), entailed IgG seroconversion rates of up to 68 % (23), a feature that was, however, not reproducible for patients receiving triple immunosuppressive therapy and being seronegative before the third vaccination (6). Already positive but low pre-vaccination IgG levels were associated with superior outcomes after a fourth dose on stable immunosuppression, leading to seroconversion rates of 42-50% (24, 25). Still, seronegative patients after repeated re-vaccination clearly represent the most vulnerable subgroup with higher risk of severe COVID19 (1, 3). This aspect is becoming increasingly important with the emergence of new viral variants, resulting in reduced neutralization capacity even in healthy individuals (26).

Our approach to withdraw MPA, followed by re-vaccination, obviously impacted all arms of immunity with a strong impact on B cell activation and differentiation, thereby boosting spike-specific antibody production. Interestingly, seroconversion was already detectable on day 7 in 34.4% of patients, as compared to only 12% of individuals receiving a third dose under MPA treatment (6), highlighting enhanced immune kinetics in the absence of anti-metabolites. Importantly, seroconversion did not depend on the type of previous vaccines since we did not observe differences between patients who received a heterologous or homologous vaccination regimen.

So far, anti-metabolites including MPA and Aza have been primarily demonstrated to impair B cell proliferation and plasmablast formation in autoimmunity (27), but to also block expansion and activation of naïve and memory B cells isolated from healthy individuals (28-30). Mechanistically, MPA inhibits IL-6-mediated STAT3-signalling, a prerequisite for plasma cell differentiation (31) and critical for their survival and immunoglobulin secretion in the bone marrow (32). To the best of our knowledge, our data for the first time verify MPA effects on B cells in an antigen-specific context, including impairment of spike-specific CD27^++^CD38^+^ plasmablast formation. With respect to the T cell compartment, our data are in line with recent studies showing that production of IFNγ, TNFα or IL-2 by polyoma-virus BK specific memory T cells or bulk mucosal-associated invariant T cells remains unaffected by MPA (33, 34). This might be related to the fact that the IL-6/STAT3 axis is selectively involved in differentiation of IL-17 secreting Th17 cells (35). As opposed to cytokine production, in vivo activation, as evidenced by Ki67 and PD-1 expression, significantly increased in vaccine-specific CD4^+^ T cells after MPA hold, suggesting that initial activation/proliferation is more sensitive to anti-metabolites as has been demonstrated for purified naïve human T cells (36). Interestingly, our finding that higher frequencies of spike-reactive T cells correlate with specific IgG levels mirrors early analyses of healthy SARS-CoV-2 vaccinees (37).

As an obvious limitation of our approach, a subgroup of patients with previous or ongoing Belatacept treatment did not benefit from MPA hold. Although patient numbers were small, our data support the notion that Belatacept efficiently inhibits vaccine responses (38) irrespective of the presence of anti-metabolites, suggesting that other approaches are needed for Belatacept-treated patients.

In summary our data provide evidence that temporal hold of MPA for 5 weeks in patients under previous CNI, MPA ± CS is a viable option to accelerate and increase vaccine efficacy in KTR, particularly given that graft function remained stable and no rejection episodes or increases in anti-HLA antibodies and dd-cfDNA plasma concentrations were observed. Our study thus highlights a potential rapid vaccination strategy for at-risk patients under standard CNI-based immunosuppression that warrants testing in larger cohorts.

## Materials and Methods

### Study protocol and participants

All participants gave written informed consent for sample collection according to the approval of the ethics committees of the Charité-Universitätsmedizin Berlin (EA2/010/21, EA4/188/20). Patient demographics are summarized in Table 1. Peripheral blood and serum samples were collected immediately before and 7±2 days after the fourth vaccination (humoral, B- and T-cell analyses) and 28-35 days (mean “day 32”) after the fourth dose (humoral analyses, HLA antibodies, assessment of dd-cfDNA).

### Serological assessment

Serological assessment was performed as previously reported (5, 6, 39). In brief, SARS-CoV-2 S1 domain specific IgG and IgA was determined by ELISA (Euroimmun). Previous or current SARS-CoV-2 infection was excluded based on medical history in combination with negativity in a SARS-CoV-2 nucleoprotein specific ELISA (Euroimmun). Samples were considered positive with OD ratios of ≥1.1 as per manufacturer’s guidelines. An OD ratio value was determined by calculating the ratio of the OD of the respective test sample over the OD of the internal calibrator provided with the ELISA kit. Virus neutralization capacity of sera was analyzed using a surrogate SARS-CoV-2 neutralization test (GenScript) with more than 30% being defined as a positive response as described previously (40, 41).

### Clinical parameters

Clinical parameters were extracted from our patient database (42).

### Erythrocyte inosine monophosphate dehydrogenase (IMPDH) measurement

Erythrocyte IMPDH activity was measured as described recently (17) as a part of clinical routine. The last available IMPDH in a steady state condition before change in MPA dose is reported in Table 1.

### dd-cfDNA Measurement

The measurement of dd-cfDNA was performed as described previously (15, 43). In brief, for each patient, 4 informative independent single nucleotide polymorphism (SNP), assays are used for which the recipient has a homozygous allelic state and the graft carries at least 1 heterozygous allele. These are selected from a predefined set of 40 SNPs. These 4 SNP assays were used to quantify the dd-cfDNA (%) concentration, defined as donor alleles/(donor alleles + recipient alleles). Results for SNPs with heterozygous graft genotypes were corrected by a factor 2. Total cfDNA was extracted from up to 8 mL of plasma collected in certified blood collection tubes (Streck Corp, Omaha, Nebraska). The measurement was performed using droplet-digital PCR. Results were corrected for extraction efficiency and cfDNA fragmentation in absolute quantification, as described previously (20). The absolute concentration of dd-cfDNA per mL of plasma was calculated by multiplying total cfDNA (copies/mL) and dd-cfDNA (%). Time dependent changes for total cfDNA and dd-cfDNA fraction (%) in the posttransplant course were assessed in a cohort of 300 KTR, as described previously (44).

### Characterization of antigen-specific B and T cells

All experiments have been performed as previously described (5, 18, 39). In brief, peripheral blood mononuclear cells (PBMCs) were isolated by density gradient centrifugation using Ficoll-Paque PLUS (GE Healthcare Bio-Sciences, Chicago, IL, USA). B cells were detected within PBMC by flow cytometry and gated as CD19^+^CD3^-^CD14^-^ among single live lymphocytes (gating strategy depicted in Supplementary Figure 1B). For flow cytometric analysis, the following fluorochrome-labeled antibodies were used: CD14 (M5E2, BD Biosciences, Franklin Lakes, NJ, USA), CD3 (UCHT1, BD), CD27 (L128, BD), CD19 (SJ25C1, BD), CD24 (ML5, BD), IgD (IA6-2, BioLegend, San Diego, CA, USA), CD38 (clone HIT2, BioLegend). Antigen-specific B cells were identified (Supplementary Figure 1 D) by double staining with recombinant purified RBD (DAGC149, Creative Diagnostics, New York, USA) conjugated to AF647 or AF488, respectively. For identification of vaccine-reactive T cells, 3-5×10^6^ PBMC were stimulated for 16 h with overlapping 15-mers covering the complete SARS-CoV-2 spike protein (1 ug/ml per peptide; JPT, Berlin, Germany). Specific CD4^+^ T helper cells were identified based on CD154 and CD137 coexpression as shown in Supplementary Figure 2. For labelling of surface markers, antibodies against CD3 (SK7, Biolegend), CD4 (SK3, BD), CD8 (SK1, Ebioscience, San Diego, CA, USA), CD45RO (UCHL1, BioLegend), CD62L (DREG-56, BioLegend) and PD1 (EH12.1, Becton Dickinson) were used. A dump channel served to exclude unwanted cells, containing CD14^+^ (M5E2, BioLegend), CD19^+^ (HIB19, BioLegend), and dead cells (fixable live/dead, BioLegend). After surface staining, cells were fixed in FACS Lysing Solution (Becton Dickinson), permeabilized in FACS Perm II Solution (Becton Dickinson) and stained intracellularly with anti-CD154 (24-31, BioLegend), anti-CD137 (4B4-1, BioLegend), anti-TNF-α (MAb11, BioLegend), anti-IFN-γ (4SB3, Ebioscience), anti-IL-2 (MQ1-17H12, BioLegend), anti-Ki67 (B56, Becton Dickinson), and anti-IL-4 (MP4-25D2, BioLegend). Data acquisition was performed using a BD FACS Fortessa X20 (BD).

### FACS data analysis and statistics

FACS data was analysed with FlowJo 10 (BD). The gating strategies for analysis of antigen-reactive B- and T cells are illustrated in Supplementary Figures 1 and 2. Co-expression of cytokines was quantified by Boolean gating in Flowjo. Statistical analysis and graph preparation was conducted in GraphPad Prism 8 (GraphPad, La Jolla, CA, USA). Normal distribution of data was assessed using the Kolmogorov-Smirnov test. Depending on normal distribution or not, a t-or Wilcoxon test was used for paired two-group comparisons. For multiple comparisons, a two-way ANOVA with Šidák’s post-test or Kruskal-Wallis test with Dunn’s post-test were chosen. For analysis of contingency tables, Fisher’s exact test was applied.

## Data Availability

All data produced in the present study are available upon reasonable request to the authors.

## Acknowledgments

The authors are grateful to all contributing patients.

## Author contribution

ES, AS FH and KB designed the study and wrote the manuscript.

HRA, ALS, ES, BP, AS, VP and CS performed experiments

ES, CH, AJ, KB, NK BZ and MC recruited patients.

LL and PG provided IMPDH data.

BO, JB and MO performed assessment of dd-cfDNA.

NL and CLDM performed HLA antibody testing.

HS, CL and BJ were responsible for serological studies.

ES, KB, KK, TD, KUE and FH supervised the work and provided funding.

---

**Supplemental Figure 1.**
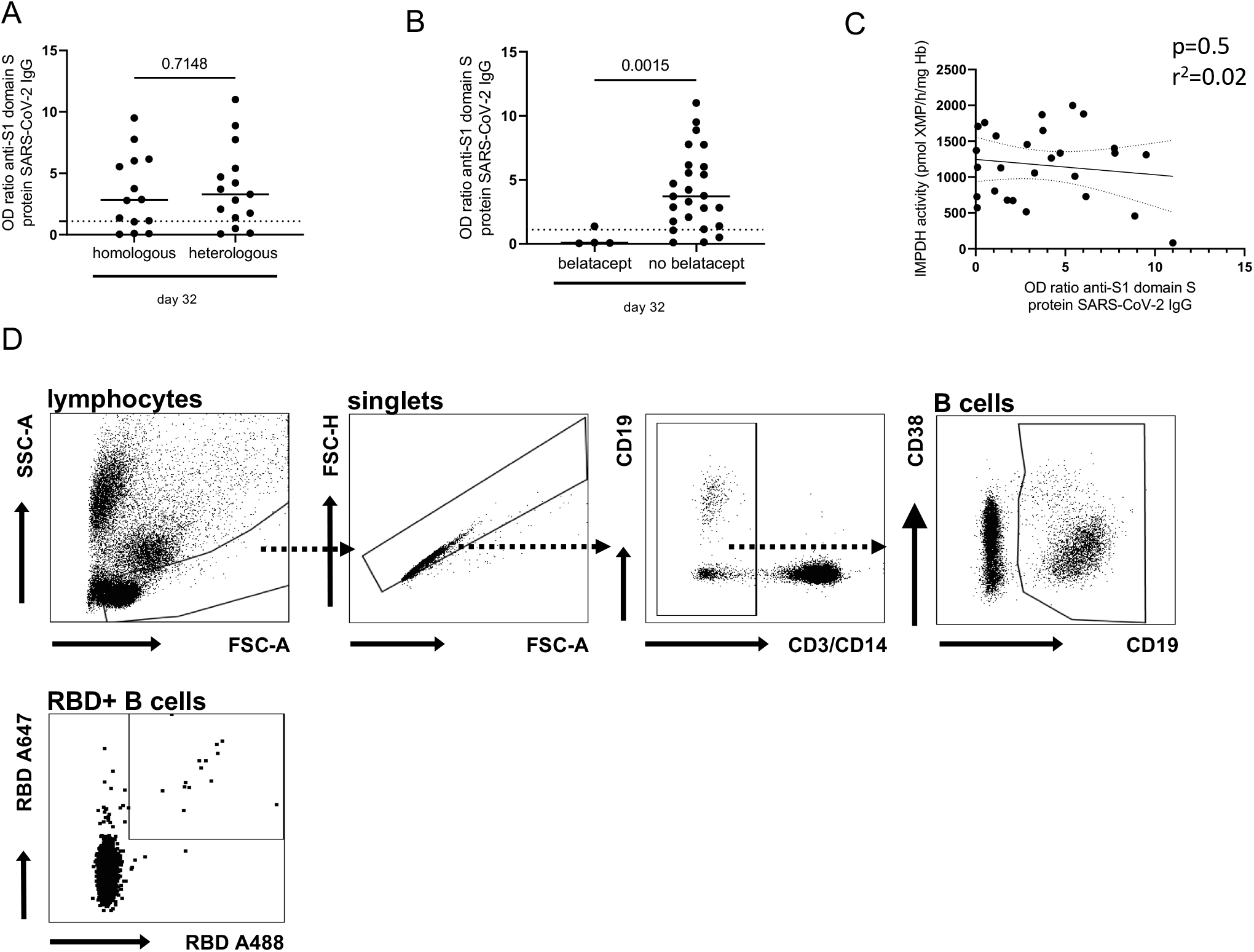
(A) Humoral vaccine-specific immune responses were assessed by ELISA for anti-spike protein S1 IgG and compared in patients with a homologous versus a heterologous previous vaccine protocol on day 32 after 4th vaccination. (B) Humoral vaccine-specific immune responses were assessed by ELISA for anti-spike protein S1 IgG and compared in patients with vs. without belatacept based immunosuppression on day 32 after 4th vaccination. The dotted lines mark the threshold for positivity (OD ratio of 1.1). (C) Correlation of inosine monophosphate dehydrogenase (IMPDH) activity in erythrocytes and anti-spike protein S1 IgG on day 32 after 4th vaccination. (D) Detection of SARS-CoV2 vaccine specific B cells. B cells in PBMCs were detect by flow cytometry. Antigen-specific B cells were identified by double staining with recombinant purified RBD conjugated to AF647 or AF488, respectively.

**Supplemental Figure 2.**
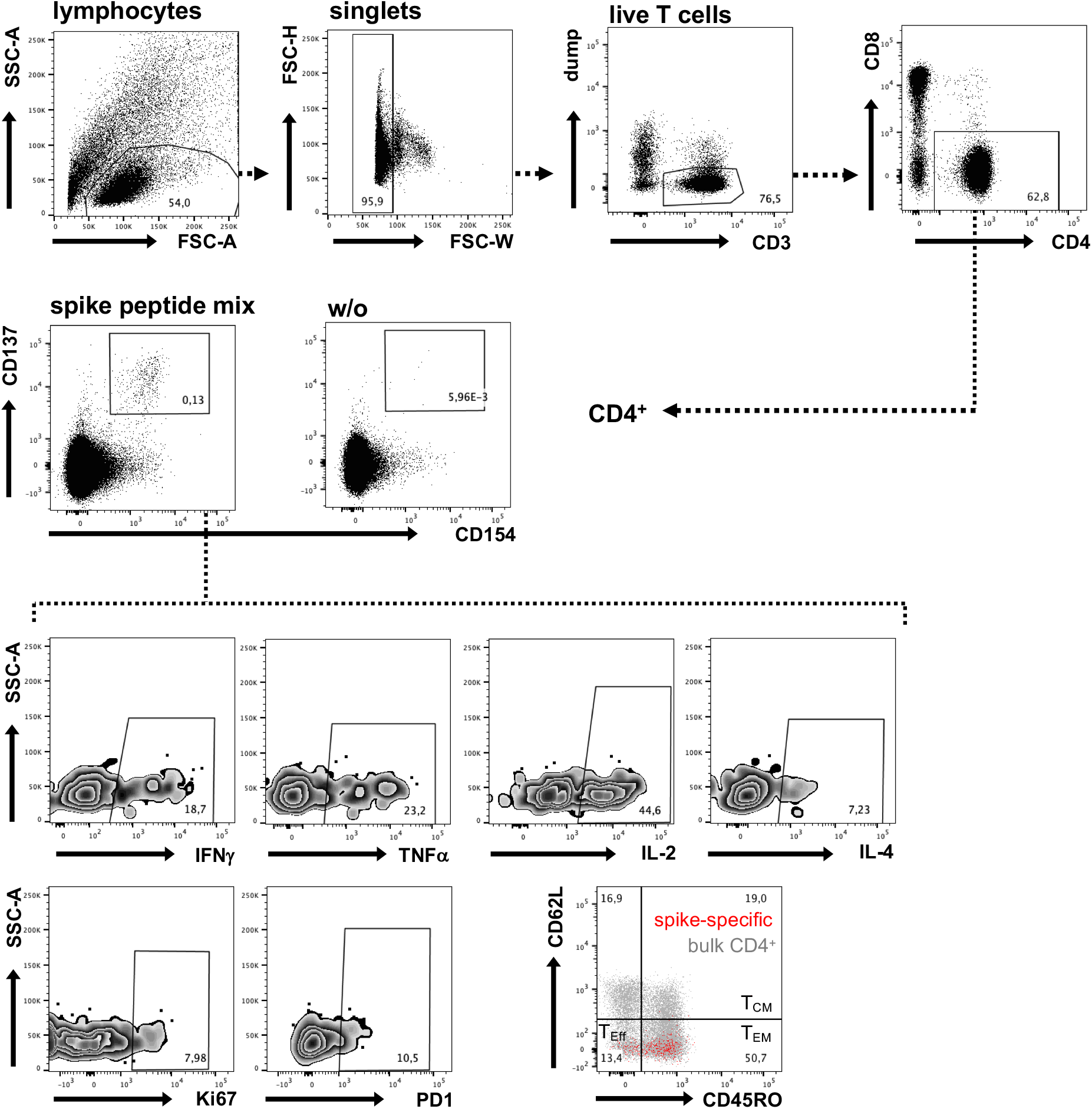
Detection of SARS-CoV-2 vaccine specific T helper cells. Human PBMC were stimulated with SARS-CoV-2 spike peptide mix or left unstimulated for 16 h. Vaccine-specific live single CD14-CD19-CD3+ (“dump” negative) CD4+ T helper cells were detected by flow cytometry based on co-expression of CD154 and CD137. Specific cells were subsequently analyzed for expression of IFNγ, TNFα, IL-2 and/or IL-4, for the in vivo induced proliferation/activation markers Ki67 and PD1, or for their memory phenotype based on CD45RO and CD62L expression (T_CM_: central memory-, T_EM_: effector memory-, T_eff_: effector T cells). Gates were set according to the respective unstimulated or unstained controls.

